# Post-vaccination SARS-COV-2 among healthcare workers in New Jersey: a genomic epidemiological study

**DOI:** 10.1101/2021.06.30.21259761

**Authors:** Barun Mathema, Liang Chen, Kar Fai Chow, Yanan Zhao, Michael C Zody, Jose R Mediavilla, Marcus H Cunningham, Kaelea Composto, Annie Lee, Dayna M Oschwald, Soren Germer, Samantha Fennessey, Kishan Patel, David Wilson, Ann Cassell, Lauren Pascual, Andrew Ip, André Corvelo, Sophia Dar, Yael Kramer, Tom Maniatis, David S Perlin, Barry N Kreiswirth

## Abstract

Emergence of SARS-CoV-2 with high transmission and immune evasion potential, the so-called Variants of Concern (VOC), is a major concern. We describe the early genomic epidemiology of SARS-CoV-2 recovered from vaccinated healthcare professionals (HCP). Our post-vaccination COVID-19 symptoms-based surveillance program among HCPs in a 17-hospital network, identified all vaccinated HCP who tested positive for COVID-19 after routine screening or after self-reporting. From 01/01/2021 to 04/30/2021, 23,687 HCP received either mRNA-1273 or BNT162b2 mRNA vaccine. All available post-vaccination SARS-CoV-2 samples and a random collection from non-vaccinated patients during the similar timeframe were subjected to VOC screening and whole genome sequencing (WGS). 62% (23,697/37,500) of HCPs received at least one vaccine dose, with 95% (22,458) fully vaccinated. We detected 138 (0.58%, 138/23,697) COVID-19 cases, 105 among partially vaccinated and 33 (0.15%, 33/22,458) among fully vaccinated. Five partially vaccinated required hospitalization, four with supplemental oxygen. VOC screening from 16 fully vaccinated HCPs identified 6 (38%) harboring N501Y and 1 (6%) with E484K polymorphisms; concurrent non-vaccinated samples was 37% (523/1404) and 20% (284/1394), respectively. There was an upward trend from January to April for E484K/Q (3% to 26%) and N501Y (1% to 49%). WGS analysis from vaccinated and non-vaccinated individuals indicated highly congruent phylogenies. We did not detect an increased frequency of any RBD/NTD polymorphism between groups (P>0.05). Our results support robust protection by vaccination, particularly among recipients of both doses. Despite VOCs accounting for over 40% of SARS-CoV-2 from fully vaccinated individuals, the genomic diversity appears to proportionally represent those among non-vaccinated populations.

**IMPORTANCE:** A number of highly effective vaccines have been developed and deployed to combat the COVID-19 pandemic. The emergence and epidemiological dominance of SARS-CoV-2 mutants, with high transmission potential and immune evasion properties, the so-called Variants of Concern (VOC), continues to be a major concern. Whether these VOCs alter the efficacy of the administered vaccines is of great concern, and a critical question to study. We describe the initial genomic epidemiology of SARS-CoV-2 recovered from vaccinated healthcare professionals and probe specifically for VOC enrichment. Our findings support the high-level of protection provided by full vaccination despite a steep increase in the prevalence of polymorphisms associated with increased transmission potential (N501Y) and immune evasion (E484K) in the non-vaccinated population. Thus, we do not find evidence of VOC enrichment among vaccinated groups. Overall, the genomic diversity of SARS-CoV-2 recovered post-vaccination appears to proportionally represent the observed viral diversity within the community.

## INTRODUCTION

Since the beginning of the pandemic in 2020, SARS-CoV-2 has infected and spread within an immunologically naïve global population (1, 2). With the application of convalescent plasma and therapeutic antibody treatment, along with highly effective vaccines, the virus is now starting to experience the immune pressure that will ultimately shape its evolutionary trajectory (3). Recent evidence indicates that SARS-CoV-2 can mutate and evade immunity and challenge the efficacy of emerging vaccines and intrinsic and extrinsic antibodies (i.e., natural by infection and antibody therapeutics) (3, 4).

The reports of mutants, known as Variants of Concerns (VOCs) that exhibit increased transmission or immune evasion, or both, have been reported in different global regions and predominant lineages have been genotyped (5). Of note, three VOCs, B.1.1.7 (recently named Alpha) (6), B.1.351 (Beta) (7), and P.1 (Gamma) (8) containing prominent mutations in the spike protein, emerged in the UK, South Africa and Brazil, respectively, and more recently, B.1.617.2 (Delta) (9) emerged in India and has spread globally. The mapping of specific mutations in the spike protein has revealed strong evidence of convergent evolution, and particularly the E484K polymorphism, which is able to evade certain monoclonal therapy and it is less responsive to neutralizing antibodies from recovered patients. This mutant shows reduced response to convalescent plasma and reported reinfections and it has been identified in discrete lineages in different geographic locations and associated with variant clones with increased incidence (4). The impact of these antibody escape mutants harboring VOCs on driving community incidence and whether they alter the efficacy of the administered vaccines is of great concern.

In this report, we describe the initial genomic epidemiology of SARS-CoV-2 recovered from vaccinated healthcare professionals (HCPs) within a large healthcare network in New Jersey. We specifically probe whether specific SARS-CoV-2 variants skew viral diversity indicative of vaccine-induced selection; and compare against a random collection of SARS-CoV-2 recovered from non-vaccinated individuals during the same time period. Although we find steep increases in variants harboring E484K and N501Y among community samples, our early genomic assessment does not indicate specific variant enrichment among post-vaccinated individuals. We find that viral genomes among vaccinated individuals largely reflect the viral diversity among non-vaccinated populations.

## RESULTS

### COVID-19 post-vaccination surveillance program: demographic and clinical characteristics of COVID-19 cases

From December 2020 to April 2021, 23,697 of 37,500 HCPs (62%) received at least one dose of an mRNA vaccine, and 22,458 (59%, 22,458/37,500) received both vaccine doses. Among vaccinated employees, 12,878 (54%) received mRNA-1273 (Moderna) and 10,819 (46%) received BNT162b2 (Pfizer/BioNTech) mRNA COVID-19 vaccine. Of the 23,697 vaccinated (single and both doses) employees, our surveillance program detected 138 (0.58%, 138/23,697) COVID-19 cases, 105 of these were among employees who received only one dose and 33 (0.15%, 33/22,458) with both doses.

Among the 138 post-vaccinated COVID-19 cases, 74 were vaccinated with the Pfizer/BioNTech vaccine and 64 were vaccinated with the Moderna preparation. Cases were reported from nine hospitals. Demographic information was not available for 17 employees. Of the remaining 121 employees, most cases were white (68%) and female (74%) with a median age of 45 (ranging from 19-77). Twenty-one employees (17%) were documented to have asymptomatic infection. Of those that were symptomatic, five individuals requiring hospitalization received only one dose and had recorded BMI of >25 (4 individuals with BMI >30) and four of these patients required supplemental oxygen. The majority of the vaccine breakthroughs, 76% (105/138), occurred from 1-113 days after the initial dose with a median of 10 days, while 23% (33/138) occurred 4-104 days after the second dose with a median of 22 days.

### Screening of post-vaccinated SARS-CoV-2 infections indicate the rise in E484K and N501Y variants

To further characterize the SARS-CoV-2 genotypes recovered from the post-vaccinated individuals, we examined the spread of key mutations underlying VOCs (*i*.*e*., B.1.1.7) in New Jersey using a high-throughput molecular beacon assay designed to screen for polymorphisms N501Y/T and E484K/Q in the RBD region (10). Among 138 HCPs, 83 swabs were available for rapid screening providing 76 genotypic results (60 from partial and 16 from fully vaccinated). There were 3 and 6 viral samples with E484K and N501Y polymorphisms respectively (Figure 1). Among 16 fully vaccinated HCPs, we found 1 (6.3%) and 6 (38%) samples with E484K and N501Y mutations, respectively.

**FIGURE 1.**
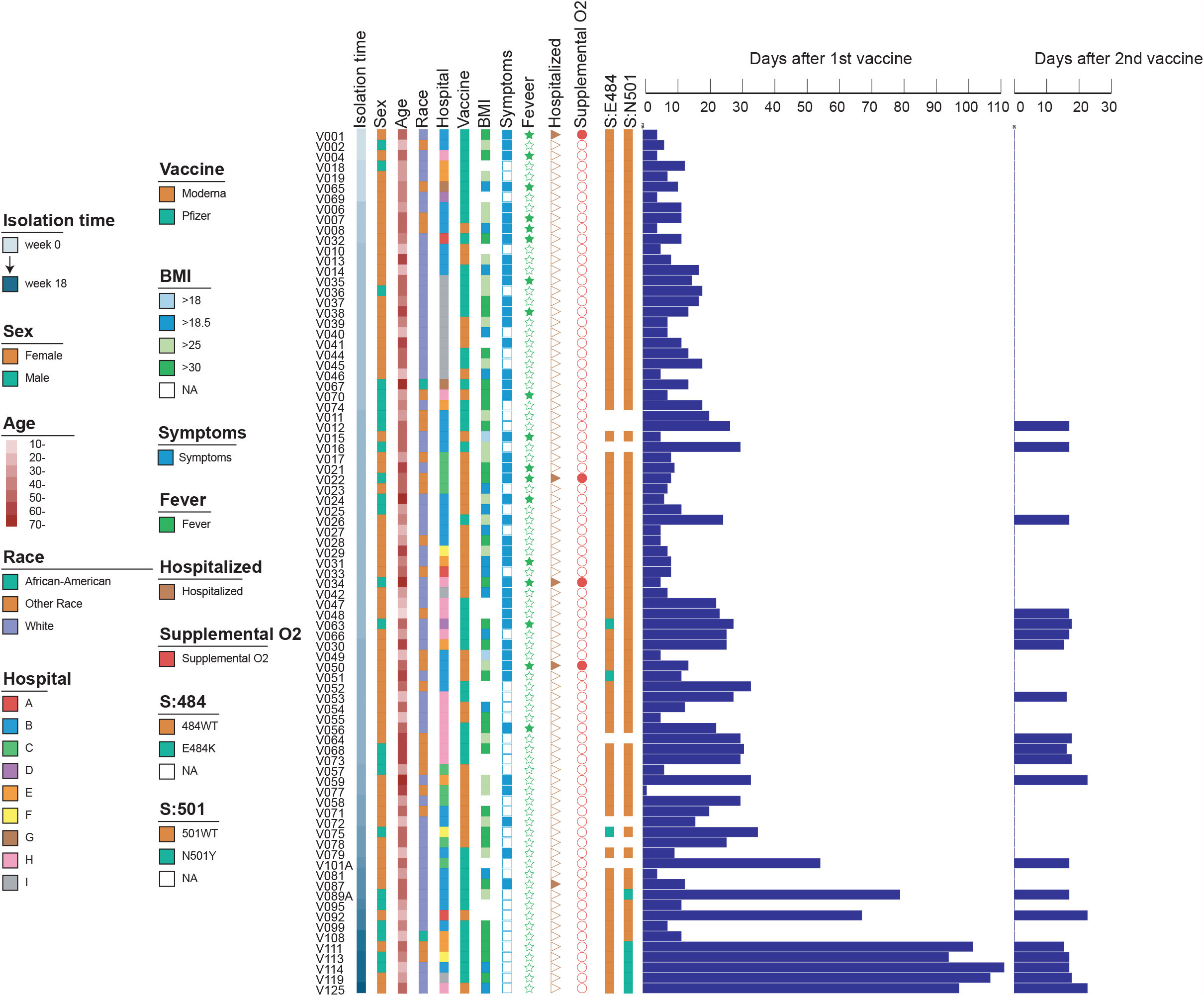
Clinical characteristics of 83 COVID-19 vaccinated HCPs and their SARS-CoV-2 S protein E484 and N501 genotypes. The samples were ordered by the time of diagnosis, with the the first case detection week denoted as “0”.

To contextualize the prevalence of E484K and N501Y mutations among the non-vaccinated population, we randomly sampled 1,404 SARS-CoV-2 positive swabs (from a total of 3,000 positive COVID-19 swabs) from January 2021 to April 2021 within our hospital network. Overall, we detected 284 E484K/Q (284/1392, 20.4%, 12 samples failed in the E484 detection in comparison with N501Y) and 523 N501Y (523/1404, 37.3%) mutants (Figure 2). The prevalence of E484K/Q and N501Y from January to April, was 3.2% to 25.7% and 1.0% to 49.3%, respectively (Figure 2).

**FIGURE 2.**
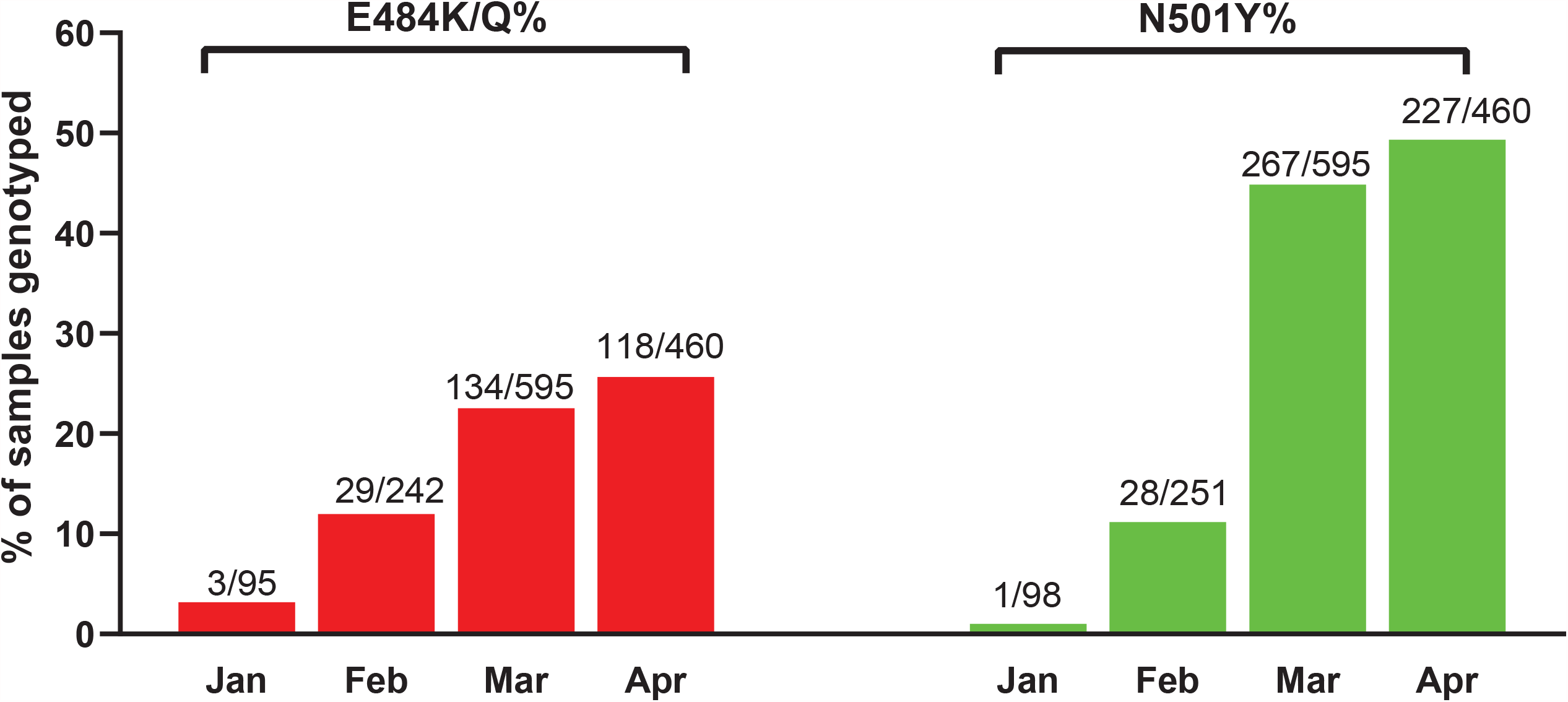
Prevalence of E484K/Q and N501Y mutants among SARS-Cov-2 samples from non-vaccinated individuals from January to April 2021.

### SARS-CoV-2 from vaccinated individuals is diverse and does not indicate predominant genotypes or genetic characteristics

To understand the population structure of SARS-CoV-2 post-vaccination, we first analyzed WGS data for 68 available SARS-CoV-2 isolates from 83 symptomatic COVID-19 cases included in this study. WGS failed for 15 samples due to high Ct values, suggestive of low viral burden. We specifically probed whether specific variants or mutations were overrepresented among SARS-CoV-2 recovered from vaccinated individuals. The 68 genomes were divided into 21 different Pangolin lineages with B.1.2 the most common (24/68, 35%) (Figure 3). We noted distinct amino acid changes or deletions at 20 sites within the N-terminal domain (NTD) or receptor-binding domain (RBD) regions of the spike protein. Mutations associated with antibody evasion - L452R, T478K, E484K and S494P - were found in 3, 1, 3 and 2 genomes, respectively. We did not detect any genomes with more than two of the four polymorphisms. The N501Y mutation was found in 4% (3/68) of the genomes, and all belonged to the VOC B.1.1.7 lineage. We recorded 3 viral samples harboring the E484K mutation, two belonged to the B.1.526 lineage first identified in New York (11, 12) and one from the R.1 lineage. Importantly, among the 9 individuals fully vaccinated (*i*.*e*., ≥14 days post-second dose) we note 1 and 2 samples harboring E484K and N501Y polymorphisms, respectfully. There were no samples from fully vaccinated HCPs with both E484K and N501Y.

**FIGURE 3.**
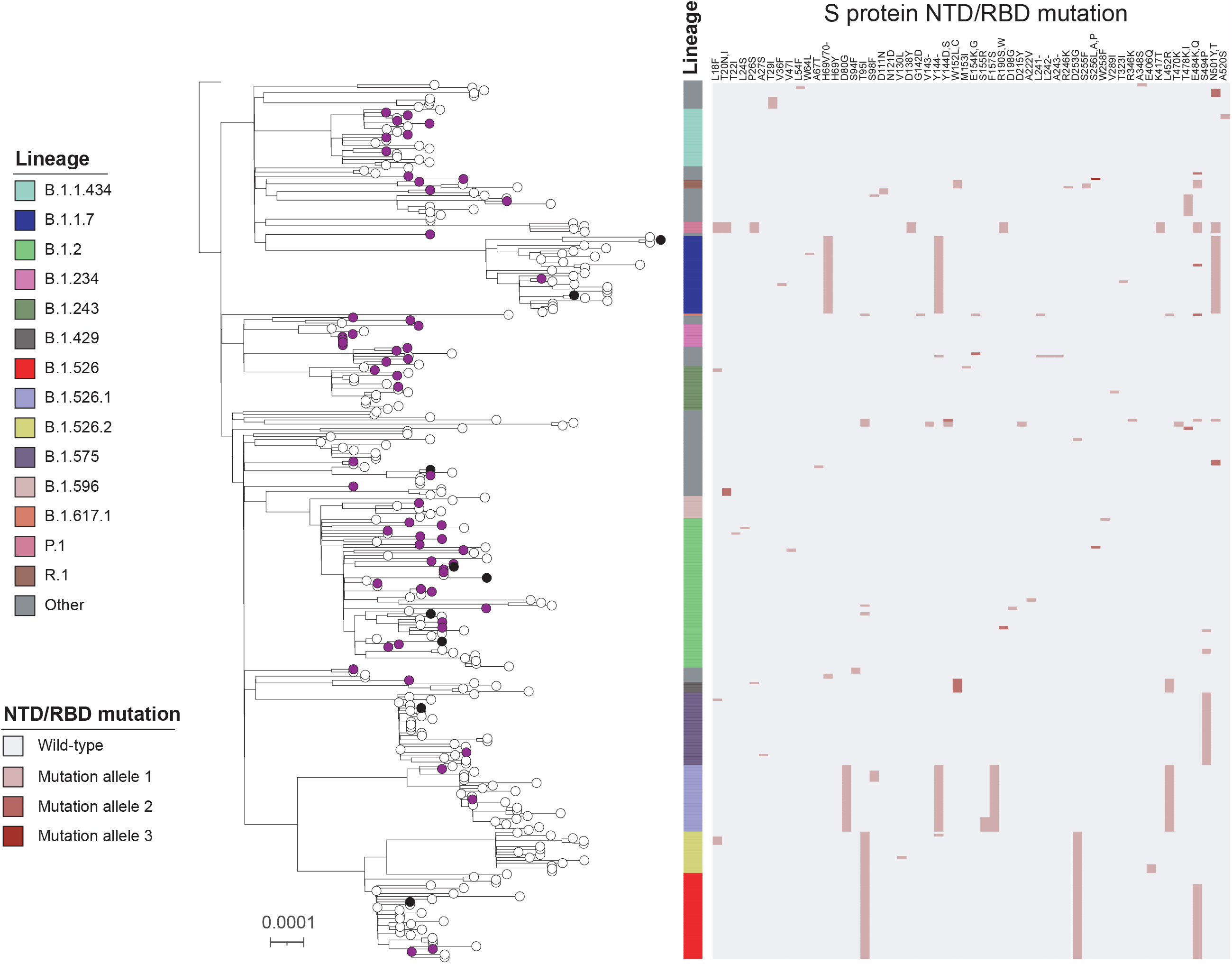
Maximum Likelihood Phylogenetic tree of 349 SARS-CoV-2 genomes isolated from vaccinated and non-vaccinated populations. The tree is rooted to the SARS-CoV-2 Wuhan-Hu-1 reference genome (NC_045512.2). The scale represents 0.0001 nucleotide substitutions per site. The vaccination conditions are color coded at the tree tips: purple, black and white tips denote single-dose, double-dose vaccine and non-vaccination, respectively. The SARS-CoV-2 Pangolin lineage is illustrated as a color bar and the S protein NTD/RBD mutations are shown as a heatmap on the right panel.

To compare viral diversity from vaccinated and non-vaccinated individuals, we randomly selected 281 SARS-CoV-2 samples recovered from non-vaccinated patients within the Hackensack Meridian Health (HMH) network during the same time period for genomic characterization by WGS (Figure 3). Among them, 38 different Pangolin lineages were identified, including 18 lineages found in the vaccinated population. B.1.526 (n=33) and its sub-lineages (B.1.526.1, n=26, and B.1.526.2, n=15) were the most common (74/281, 26.3%), followed by B.1.2 (35, 12.5%), B.1.1.7 (28, 10%) and B.1.575 (26, 9.3%). In addition, 5 P.1 (20J/501Y.V3) and 1 B.1.617.1 (Kappa) were found among the 281 samples. Mutations or deletions were detected in 54 sites within the S protein RBD or NTD region, including 18 sites found in the vaccinated samples. L452R, T478K/I, E484K/Q and S494P mutations were identified in 32, 8, 40 and 29 genomes, respectively. Compared with the non-vaccinated samples, we did not detect a significant increase in the frequency of any RBD/NTD mutations or deletions among the vaccinated group (P > 0.05).

## DISCUSSION

Rapid expansion and contraction of SARS-CoV-2 populations coupled with increasing anti-CoV-2 immunity dynamics have contributed to pandemic phases accompanied by emergent genomic signatures (1, 13). Extensive polymorphisms in the spike protein have been documented, including N501Y and E484K mutations that are particularly concerning (14). In this report, we present early genomic analysis of SARS-CoV-2 from an ongoing post-vaccination COVID-19 surveillance program among healthcare workers in New Jersey.

We detected SARS-CoV-2 in 33 (0.15%) HCPs who were fully vaccinated, none of whom required hospitalization. All symptomatic cases were noted among partially vaccinated HCPs and were self-limiting with only 5 requiring hospitalization. Although our passive surveillance system was intended to capture the HCP vaccinated population who self-reported symptoms or who were exposed to a COVID-19 positive case, and not able to report true prevalence of infections (i.e., mild or asymptomatic infections may be missed), our findings are consistent with emerging reports of incident post-vaccination SARS-CoV-2 infections (15-18). A recent multicenter prospective cohort study in England found vaccine effectiveness upwards of 85% (19). Our results support high levels of protection afforded by vaccination, particularly among recipients of both doses (9, 15, 20-24). The majority of cases in our study were among partially vaccinated individuals, emphasizing the importance of personal protective measures, and susceptibility to infection particularly until fully vaccinated (19, 24-27).

Steep transmission gradients reported locally and regionally have been partly attributed to VOCs, including B.1.1.7 in Europe and U.S., P.1 in Brazil, B.1.351 in South Africa, B.1.617.2 in India and B.1.526 in New York State (6-8, 11, 28). Although the pathogen characteristics attributable to the spread of these variants at the population-level are difficult to assess, mathematical modeling, genomic and experimental studies have demonstrated increased transmissibility and immune evasion properties, including reduced vaccine-induced antibody neutralization *in vitro* (2, 3, 6, 14). Whether the rollout of vaccination would offer selective advantage to VOCs is a critical question to determine.

A recent study from Washington State found that all SARS-CoV-2 vaccine breakthroughs were VOCs. Of significance, those VOCs harboring immune evasion properties (B.1.351, B.1.427, B.1.429, and P1) were enriched compared to B.1.1.7 lineages among vaccine breakthrough cases when compared to cases in the general population (29). Other outbreaks have been reported among vaccinated individuals implicating SARS-CoV-2 strains harboring E484K, a noted immune evasion conferring polymorphism (20). A recent study where two VOCs (B.1.351 and B.1.1.7) dominate the viral population, indicate reduced vaccine effectiveness against both variants at specific time windows. Reduced effectiveness against B.1.351 at least 7 days after the second BNT162b2 dose, and against B.1.1.7 between 2 weeks after the first dose and 6 days after the second dose were reported (24). However, among fully vaccinated HCPs the VOC proportions between cases and controls were comparable, consistent with our findings. These studies reinforce the need to fully vaccinate to achieve high levels of protection afforded by vaccination (9).

In our study, despite an increasing trend in the proportion of N501Y and E484K variants in the overall population (Figure 2), we do not find early evidence of genotypic enrichment of polymorphisms within the NTD or RBD region of the spike protein gene among our fully vaccinated HCPs. Although 44% of the strains recovered from fully vaccinated HCPs harbored mutations of concern (6 N501Y and 1 E484K), they were similar in proportion to strains circulating in the community during the same time period. Our WGS analysis suggests N501Y harboring strains belong to the B1.1.7 lineage and the E484K mutants to the B.1.526 lineage, both highly prevalent lineages in Northeastern US (12, 30). All N501Y mutants were detected among fully vaccinated HCPs, half of which failed to generate WGS data due to higher Ct values, a likely consequence of vaccination (31).

A recent report showed that B.1.1.7 upregulates key innate immune antagonists likely increasing the transmission potential of this VOC (32). Interestingly, mRNA vaccines have shown high levels of efficacy against B.1.1.7 infections (22). Overall, the genomic diversity of SARS-CoV-2 recovered post-vaccination appears to proportionally represent the observed viral diversity within the community. We acknowledge that our estimates may be skewed by under-sampling of asymptomatic infections among fully vaccinated individuals (33), and likely affected by test seeking behavioral bias (24). However, our results are consistent with those from a large nationwide surveillance program of COVID-19 vaccine breakthrough infections that found the proportion of reported vaccine breakthrough infections attributed to VOCs to be similar to the proportion of these variants circulating throughout the United States (27).

There is mounting evidence that currently available vaccines, including those used in our population, are highly effective against VOCs. The reports of VOC enrichment among post-vaccinated individuals, despite small sample sizes, warrant surveillance. Our study findings are preliminary yet highlights the importance of full vaccination against circulating variants, including VOCs, and the need to continue SARS-CoV-2 genomic surveillance as vaccination coverage expands, pathogen evolves and immunity wanes.

## METHODS

### Post-vaccination COVID-19 surveillance program

Since the first mRNA vaccines were administered in December 2020 among the 37,500 HMH employees, we initiated a symptoms-based surveillance program to identify and report the status of each vaccinated person in the Electronic Medical Record (EMR). This regularly updated list was cross-referenced daily against the results of all SARS-Cov-2 nucleic acid amplification tests (NAAT) in the HMH network. As a result, we were able to identify all vaccinated employees who tested positive for COVID-19 by NAAT after routine screening or after reporting out sick. Non-team members who were vaccinated at our mass-vaccination site or by physician report (and were thus in the HMH EMR) and subsequently tested positive for COVID-19 by NAAT were also similarly identified. The study was approved by Hackensack Meridian Health Institutional Review Board (IRB).

### Whole genome sequencing and phylogenetic analysis

SARS-CoV-2 targeted assay libraries were prepared using the Molecular Loop Viral RNA Target Capture Kit (Molecular Loop) or QIAseq FX DNA Library UDI Kit (QIAGEN), in accordance with manufacturer’s recommendations. Final libraries were quantified using fluorescent-based assays including PicoGreen (Life Technologies), Qubit Fluorometer (Invitrogen), and Fragment Analyzer (Advanced Analytics). Final libraries were sequenced on a NovaSeq 6000 sequencer (v1 chemistry) with 2×150bp.

For QIAseq libraries, read pairs that did not contain a single 19bp seed k-mer in common with the SARS-CoV-2 genome reference (NC_045512.2) were discarded. Adapter sequences and low-quality bases (Q < 20) were trimmed from the 3’ end of the remaining reads, using Cutadapt v2.10 (34). Sequences corresponding to the amplicon primers were also clipped from the 5’ end of reads. Processed read pairs were then aligned to the SARS-CoV-2 reference genome, using BWA-MEM v0.7.17 (35) and only read pairs with at least one alignment spanning a minimum of 50 bp in the reference were kept.

For Molecular Loop libraries, the two 5bp UMIs located at the 5’ end of each mate were first clipped and combined into a single UMI tag. Additional 25 bp, corresponding to the molecular inversion probes, were also clipped from the 5’ end of each read. Next, read pairs that did not contain a single 19bp seed k-mer in common with the SARS-CoV-2 genome reference (NC_045512.2) were discarded and adapter sequences and low-quality bases (Q < 20) were trimmed from the 3’ end of the remaining reads, using Cutadapt v2.10 (34). Processed reads pairs were then merged using NGmerge v0.2 (36), allowing for dovetailed alignments. The resulting single end reads were mapped against the SARS-CoV-2 genome reference using BWA-MEM v0.7.17 (35) and the resulting alignments filtered using the following criteria: 1) reference span ≥ 50bp, 2) quality ≥ 60, and 3) maximum soft-clip length on either end ≤ 30bp. Next, reads representing the same original molecule were identified based on their shared UMI and alignment position and used to draw the molecule consensus sequence, taking into account base quality scores. Molecule sequences were then realigned to the SARS-CoV-2 genome reference using BWA-MEM v0.7.17 (35). Finally, genome sequences were determined either by read (QIAseq) or molecule (Molecular Loop) alignment pileup consensus calling with a minimum support of 5 reads/molecules.

The resulting SARS-CoV-2 viral genome sequences were characterized by Nextclade CLI (v1) (https://clades.nextstrain.org/) to assign Nextstrain clades (37). SARS-CoV-2 lineages were determined using Pangolin v3.1.3 (https://github.com/cov-lineages/pangolin) and GISAID clades were determined based upon the clade-specific marker variants from https://www.gisaid.org (38). The genomes were aligned using nextalign v1.0.0 (https://github.com/neherlab/nextalign), using default setting. A maximum likelihood phylogenetic tree was constructed using IQ-TREE v2.1.2 (39) with automatic model selection and 1000-bootstrap replicates. The resulting tree was annotated using ITOL v6 (40).

### Population sampling and screening of SARS-CoV-2

SARS-CoV-2 positive swabs collected from seven HMH network hospitals were shipped to CDI on a weekly basis and stored at −80°C. 1,404 swabs collected during the same time frame as the vaccinated samples were randomly selected and subjected to a molecular beacon-based real-time asymmetric PCR and melting curve analysis to identify the SARS-CoV-2 E484K/Q and N501Y mutations, as previously described (10). In brief, 50 µl aliquots of SARS-CoV-2 swab specimens were treated by proteinase K and heated at 95°C for 5 min. 5ul of heat inactivated sample was used as template for the asymmetric RT-PCR testing on a Mic Real Time PCR Cycler (Bio Molecular Systems, software micPCRv2.8.13). Based on the different melting profiles (Tm) of E484 and N501 molecular beacons in each sample, the E484K and N501Y mutations were determined.

## Data Availability

The SARS-CoV-2 genomes sequenced in this study were deposited in GISAID (www.gisaid.org). Sequences can be accessed by searching records from both the originating lab at Hackensack Medical Center and the submitting lab at the New York Genome Center.

## Statistical analysis

Fisher’s exact or chi-square tests, as appropriate, were used to examine whether the spike protein NTD/RBD mutations were enriched in the vaccinated HCPs (n=68) in comparison with the non-vaccinated patients (n=281). SPSS, version 17.0 (IBM Corp., Armonk, NY) was used for statistical analyses and two-tailed P values ≤0.05 were considered statistically significant.

